# Association of infections and autoimmune conditions with cognition: a study using self-reported conditions and identifying a novel plasma biomarker

**DOI:** 10.64898/2026.02.13.26346282

**Authors:** Patrick S. Slama, Adam R. Macbale, Bruno M. Jedynak

**Affiliations:** Independent researcher; Department of Mathematics + Statistics, Portland State University

**Keywords:** infections, autoimmune, dementia, cognitive impairment, plasma biomarkers, SIMOA, NULISA, WRAP, Trails B

## Abstract

a

**BACKGROUND:** Over the past couple of decades, the role of infections, as well as the involvement of the immune system, have been highlighted in the development of dementia.

**METHOD:** Data from the Wisconsin Registry for Alzheimer’s Prevention cohort were utilized for the analysis. A history of medical conditions was searched across the cohort, and known infections and autoimmune conditions were recorded for each participant. These conditions were then compared with the diagnosis and cognitive performances of each participant. Furthermore, plasma markers were analyzed using two different protein quantification methods.

**RESULTS:** Our analysis revealed poorer cognitive performances among participants with listed medical conditions. In plasma samples, Ab42/ICAM1 was identified as a protein ratio with significant variation across condition statuses.

**DISCUSSION:** Our study confirmed that infections and autoimmune conditions contribute to cognitive decline. Ab42/ICAM1 was identified as a relevant marker.

## b Background

Various environmental factors have been proposed as influencing the progression of Alzheimer’s disease and related dementia (ADRD). Over the past years, multiple cohort studies have thus highlighted a correlation between infections and the development of dementia. Using time series data from FinnGen, Levine and coworkers identified 45 viral exposures which were significantly associated with an increased risk of neurodegenerative disease [1]. In the Northern Manhattan study, Cytomegalovirus (CMV) and HSV-2 serologies were associated with impaired executive function, whereas *Chlamydia pneumoniae* serology was associated with impaired performance on language testing [2]. Letenneur and coworkers evidenced a strong correlation between the reactivation of HSV seropositivity and incident ADRC (DSMIII-R criteria for dementia) [3]. In their study of 575 adults from the Baltimore ECA cohort, Wennberg et al. found that a higher global burden of common infections (positive antibody tests to HSV-1, CMV, EBV, VZV, and *T. gondii*) was significantly associated with poorer cognitive performances [4]. Moreover, a study on 33,000 subjects in Taiwan showed that the risk of developing dementia was 2.56 times higher among those with an active HSV infection [5]. In a historical cohort study of nearly 1 million UK adults aged 65 and older, cases of common infections resulting in hospitalization, such as sepsis, pneumonia or urinary tract infections, were strongly associated with an increased long-term risk of incident dementia [6]. In [7], the authors evidence relationships between common adult infections and vaccinations with the risk of developing ADRC (ICD-9-CM diagnostic code 331.0).

Other studies examined whether eliminating infections had a positive impact on cognition or dementia rates. A reduced incidence of dementia after vaccination against shingles was thus recently evidenced across various analyses [8, 9]. Several vaccinations, including Tdap/Td, Herpes Zoster, and pneumococcal, were moreover associated with a reduced risk for developing ADRD [10]. In addition, analyzing medical history for a million Spanish people suggested a significantly increased risk of having ADRD when affected by autoimmune conditions such as lupus, pernicious anemia or rheumatoid arthritis [11].

Gate et al. [12] identified a specific immune signature in Alzheimer’s patients: proinflammatory *CD*8^+^*T_EMRA_*cells which clonally expand in the cerebrospinal fluid. Laurent et al. [13] demonstrated that tau pathology triggers the infiltration of CD8+ T cells into the hippocampus, which significantly contributed to cognitive decline. Furthermore, recent studies have highlighted a correlation of dementia with immune markers, including interferons, interleukins and T-cell-specific markers, as well as neuronal and glial markers, such as Neurofilament Light (NFL), Glial Fibrillary Acidic Protein (GFAP) and Chitinase-3 like-1 (CHI3L1) [14, 15]. Bu et al. [16] showed that higher infection burden was linked to significantly elevated levels of serum beta-amyloid proteins (*Aβ*40, *Aβ*42, and total *Aβ*), and proinflammatory cytokines such as TNF-*α* and IL-6.

Taken together, there is compelling evidence to suggest the possibility that infections contribute to ADRD risk through sustained immune activation, rather than merely serving as correlates of late-life cognitive decline. These aforementioned findings serve to reframe cognitive decline as, at least in part, an immune-driven process, whereby cumulative infectious exposure amplifies cognitive deterioration. Motivated by this framework, we collected medical history of viral, bacterial, and autoimmune conditions (VBA) in participants of the Wisconsin Registry for Alzheimer’s Prevention (WRAP). We studied whether this history was associated with cognitive status and cognitive impairment across a battery of neuropsychological tests. We further examined whether these associations were reflected in plasma biomarkers as measured by Single Molecule Array (SIMOA) as well as the NUcleic acid Linked Immuno-Sandwich assay (NULISA). Based on the NULISA data, we proposed a combined marker that is significantly different across VBA levels and across cognitive diagnosis levels.

## c Methods

### c.1 Cohort, medical history, clinical, imaging, and genetic data

The Wisconsin Registry for Alzheimer’s Prevention (WRAP) is one of the world’s largest and longest-running cohort studies focused on identifying the early biological and lifestyle factors that predict AD. The study is “risk-enriched,” with about 73% of participants having a parental history of the disease. This study has as a primary objective to distinguish normal cognitive aging from preclinical AD and to determine how genetics, medical history and biomarkers influence cognitive trajectories. With a long follow-up time and detailed cognitive testing, it provides a privileged context to study how a history of infections and autoimmune conditions leads to cognitive impairment.

We used the data from the WRAP November 2024 freeze cohort [17]. Since the existing literature mentions possible involvement of viruses, bacteria, and autoimmunity in the development of dementia, we chose to consider all reported viral and bacterial infections, along with reported autoimmune conditions, which we collectively designate as VBA (Viral, Bacterial, and Autoimmune). We thus performed a word search for a history of viral and bacterial infections and autoimmune diseases in the medical history spreadsheets of the WRAP cohort. Table 1 provides the list of conditions identified, along with their count and category. A participant will be denoted VBA+ if they have reported at least one of the conditions listed in this table, and VBA- otherwise. Note that this condition could have appeared before the beginning of the study or during the study, up to the last visit.

**Table 1:** List of documented health conditions, their categories, and respective counts. Category abbreviations: V, viral, B, bacterial, BV, predominantly bacterial+viral (counted as bacterial), VB, predominantly viral+bacterial (counted as viral), AI, autoimmune, R, respiratory.

| Condition | Count | Categories | Condition | Count | Categories |
| --- | --- | --- | --- | --- | --- |
| Bronchitis | 95 | V-R | Chronic Cough | 3 | VB-R |
| Lyme Disease | 53 | B | Pleurisy | 3 | VB |
| Pneumonia | 51 | BV-R | Eye Infection | 2 | B |
| Meningitis | 33 | VB | Autoimmune Disease | 2 | AI |
| Hepatitis | 20 | V | Anaplasmosis | 2 | B |
| Lupus | 16 | AI | Sjögren’s Syndrome | 2 | AI |
| Grave’s Disease | 15 | AI | Autoimmune hepatitis | 1 | AI |
| Herpes | 10 | V | Lung Infection | 1 | B-R |
| Polio | 8 | V | Intestinal Virus | 1 | V |
| Influenza | 6 | V-R | Hepatitis Arthritis Syndrome | 1 | V |
| Guillain–Barré Syndrome | 5 | AI | Hearing Loss due to Ear Infections | 1 | B |
| Syphilis | 5 | B | Hashimoto’s Disease | 1 | AI |
| COVID | 4 | V-R | Encephalitis | 1 | BV |
| Mononucleosis | 3 | V | Epstein–Barr Virus | 1 | V |
| Kidney Infection | 3 | B | Respiratory Issues during Colds | 1 | VB-R |
| Crohn’s Disease | 3 | AI |  |  |  |

The medical questionnaire given to the participants in WRAP did not specifically target VBA conditions. This questionnaire contains a list of questions starting with: “Have you experienced or has a health care provider ever told you that you have any of the following conditions?” Among the conditions, Meningitis, Chronic Bronchitis, lung disease, liver disease, and Lyme’s disease were explicitly mentioned. Other questions had to be answered as free text. Note that diseases such as Influenza or COVID-19, which have a high incidence in the general population, appear to have been under-reported. This could be explained by the absence of specific questions about these diseases in the questionnaire.

We defined “positive cognitive diagnosis” (C+) for a participant which is clinically classified with dementia, clinical mild cognitive impairment (MCI), or cognitively unimpaired-declining (*i.e.*, subclinical impairment) at their last study visit, as previously defined [18, 19, 20]. We excluded participants with the “impaired but not MCI” diagnosis, corresponding to participants having a low performance that is not due to a dementing disorder, *e.g.* learning disability or transient cause for impairment. The participants with a diagnosis of “cognitively unimpaired-stable” at their last study visit were defined as C-.

We define Amyloid positive (A+) for participants whose Pittsburgh Compound B (PiB) index value at the time of their last visit was equal to or exceeded the threshold centiloid value of 17 [21, 22]. Finally, we define as ApoE4 positive (*ApoE*4^+^) participants which have at least one ApoE4 variant, as determined by competitive allele-specific polymerase chain reaction-based genotyping assays (LGC Genomics, Beverly, MA) [17].

### c.2 Cognitive tests

Participants were submitted to various cognitive tests throughout the study, including the following: Wechsler Abbreviated Scale of Intelligence; Wide Range Achievement Test—3rd Edition Reading subtest; Rey Auditory Verbal Learning Test; Boston Naming Test—2nd Edition; Clock Drawing Test; Controlled Oral Word Association Test; Wechsler Adult Scale of Intelligence-III: Digit Span; Letter-Number Sequencing subtests; Trail Making Tests; Stroop Neuropsychological Screening Test; Brief Visuospatial Memory Test—Revised; Wechsler Adult Intelligence Scale—Revised Digit Symbol subtest; Wechsler Memory Scale—Revised Logical Memory subtest; Mini–Mental State Examination; Animal Fluency; Speech samples: open-ended interview questions and picture description; and COGSTATE: Groton Maze, One Card Learning, Paired Associates, One | Two-Back. The detailed protocol for each test can be found in [17], together with a reference.

### c.3 Protein quantifications

Plasma proteins were measured using the SIMOA platform [23] at the University of Wisconsin-Madison Alzheimer’s Disease Research Center. Four-plex assays for Ab40, Ab42, GFAP, and NFL were run on the Neurology 4-plex kit, while pTau-217 was measured with an AlzPath kit. The functional lower limits of quantification (LLOQ) were: Ab40: 4.08 pg/mL; Ab42: 1.51 pg/mL; GFAP: 11.6 pg/mL; NFL: 1.6 pg/mL; pTau-217: 0.0098 pg/mL. A comprehensive panel of 120 proteins was obtained using the NULISA approach [24].

The NULISA worklow achieves high sensitivity by combining immuno-complex formation with two successive captures by paramagnetic beads, resulting in a ≥ 10,000-fold improvement in limit of detection (LOD) over traditional proximity ligation assays. It was carried out using a preprogrammed MicroLab STARlet liquid handler, integrated with a KingFisher Presto magnetic bead processor and a BioTek ELx405 microplate washer, as already described [22, 24].

### c.4 Selection of the plasma proteins

All available proteins from the SIMOA assay were analyzed. In the NULISA workflow, from a total of over 100 available markers, 28 different proteins were considered in our analysis. These proteins were selected as being mentioned, directly or indirectly, in the dementia literature (p-Tau, Ab40, Ab42, ACHE, APOE, BACE1, FABP3, GFAP, NFL, CHI3L1, TREM2, VCAM1, VEGFD) [14, 15, 25], for their role in the immune response (ICAM1, IL2, IL7, IL33, TNF, IFNG, NRGN), or as displaying a conserved trend across VBA levels (Ab42, APOE, SNCA, CNTN2, NPTX2; this study, see Suppl. Figure 5). A full list of the proteins is provided as Table 6.

### c.5 Statistical methods

The statistical methods used in this research are cross-sectional. For each participant in the longitudinal WRAP cohort and each statistical test, the “last visit”, defined as the last visit at which the biomarker of interest is available, was selected. VBA was treated as a binary variable taking values VBA+ or VBA-. Diagnosis was also treated as a binary variable, taking values D+ or D- and similarly for ApoE4 and amyloid positivity. Trails B time was treated as a continuous variable, as well as the protein concentrations in the SIMOA and NULISA assays. We analyzed correlations between pairs of variables. In the case of two binary variables, we used Fisher’s exact test (FET). In the case of a binary and a continuous variable, we used the Wilcoxon rank-sum test (WRST, also named Mann–Whitney U test). Where the WRST returned significant results, we performed a follow up regression analysis to control for the effects of covariates using the Freedman-Lane (FL) permutation regression procedure [26]. In the case of two continuous random variables, we used the Spearman correlation coefficient and the Spearman Correlation Test (SCT). For each test, we selected the larger sample size for which the variables of interest were available.

When performing regression and logistic regression with covariates, the p-value was computed using the Normal approximations when the criteria for the Normality of the residuals were met. In cases where they were not met, we used the FL permutation test, or its generalized implementation in the GLMperm package in the case of logistic regression [27]. The corresponding ANOVA tables are presented in the Supplementary material. All the computations were performed using R Statistical Software (version 4.5.1) [28].

### c.6 Statistics on protein counts

Correlations between detected protein amounts NULISA Protein Quantifications (NPQ) were evaluated using Spearman correlation, with *ρ*(*V BA*−) designating the correlation restricted to VBAparticipants and *ρ*(*V BA*+) designating that among VBA+ participants.

## d Results

### d.1 Correlation of VBA with diagnosis and cognition

Medical history information for the 1776 participants was analyzed across the study. A list combining 31 infections and auto-immune conditions (VBA) was retained, with a total count of 353 such health conditions (Table 1; see *Methods*). Bronchitis was the disease observed with the highest frequency, followed by Lyme disease and pneumonia. Lupus (Systemic lupus erythematosus) and Grave’s disease were the most frequent auto-immune conditions, with 16 and 15 cases reported, respectively.

We first analyzed a relationship between the presence of VBA and a positive clinical diagnosis for Alzheimer’s disease. While the proportion of study participants with a positive diagnosis was 24.2% among those with no listed VBA (*P* [*D* + |*V BA*−]=0.242), this proportion increased to 30.5% for those with 1 or more VBA reported (Figure 1, left, and Table 2).

**Figure 1:**
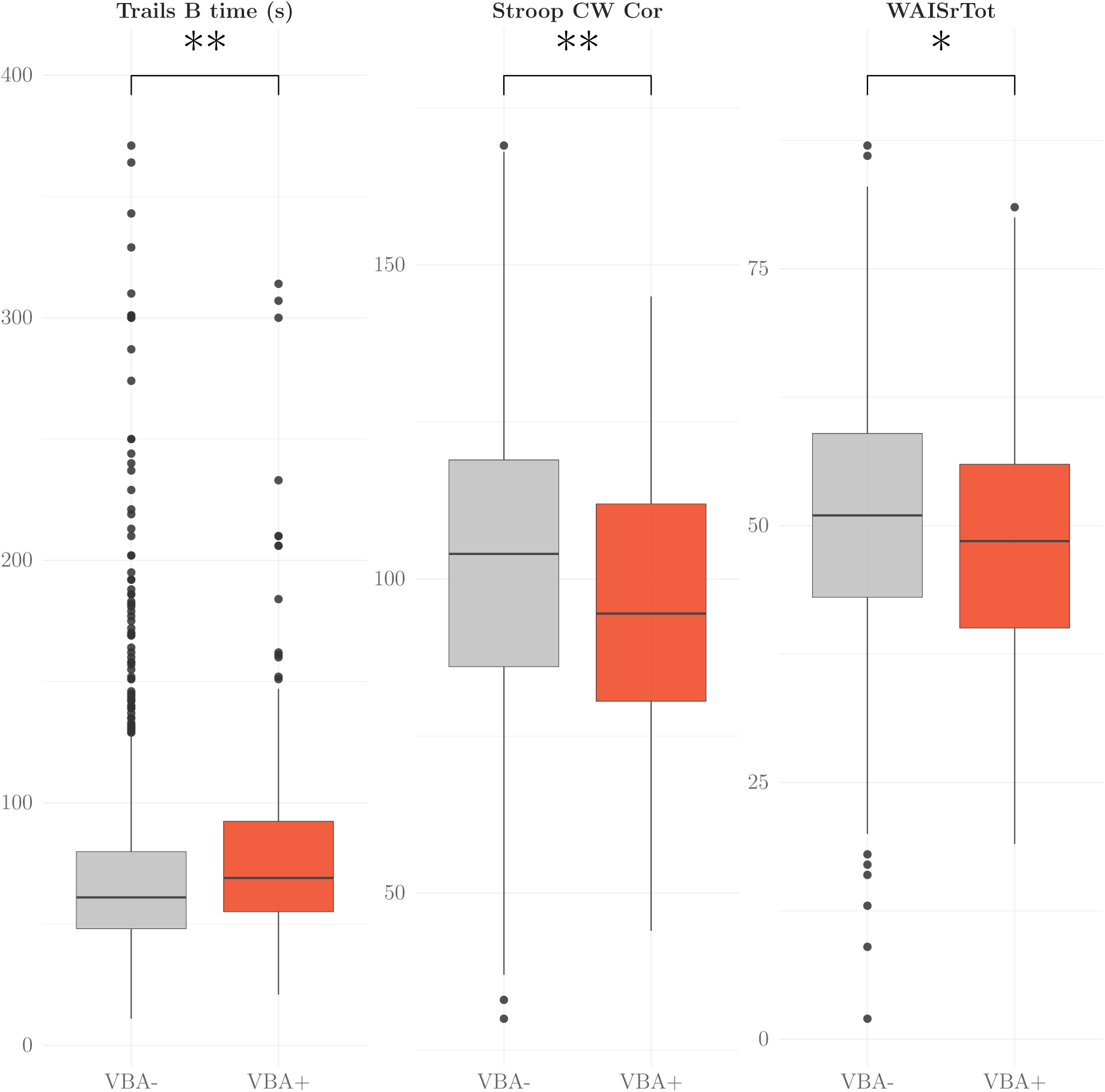
Boxplots (median, interquartile range, and 1.5×IQR whiskers) for Trails B time [lower is better], Stroop Test Color–Word Corrected (StroopCW Cor) [higher is better], and Wechsler Adult Intelligence Scale–Revised Digit Symbol Total Score (WAISrTot) [higher is better] as a function of VBA status. Significance adjusted for covariates using regression analysis (see Table 7); ∗: *p <* 5 × 10*^−^*^2^, ∗∗: *p <* 5 × 10*^−^*^3^.

**Table 2:** Detailed characteristics for the participants, based on their VBA level (no VBA documented (VBA-) or 1 or more documented (VBA+)).

|  | level |  | p |
| --- | --- | --- | --- |
|  | VBA- | VBA+ |  |
| Participant characteristics for the Diagnosis analysis |  |  |  |
| N (%) | 1491 (84.0) | 285 (16.0) |  |
| Cognitive status / diagnosis (%) |  |  | 0.030 |
| Unimpaired – stable | 1130 (75.8) | 198 (69.5) |  |
| MCI, CU-D, or Dementia | 361 (24.2) | 87 (30.5) |  |
| Age (last visit) (mean (SD)) | 65.54 (9.13) | 69.39 (8.11) | < 0.001 |
| Sex (%) |  |  | < 0.001 |
| F | 1035 (69.4) | 221 (77.5) |  |
| M | 456 (30.6) | 64 (22.5) |  |
| ApoE4 status (%) |  |  | 0.005 |
| 0 ε4 alleles | 787 (58.7) | 184 (69.2) |  |
| 1 or 2 ε4 alleles | 553 (41.3) | 82 (30.8) |  |
| Education, years (mean (SD)) | 15.87 (2.82) | 16.15 (2.92) | 0.132 |
| Race (%) |  |  | 0.086 |
| Black | 269 (18.0) | 40 (14.0) |  |
| Non White/Non Black | 35 (2.3) | 3 (1.1) |  |
| White | 1187 (79.6) | 242 (84.9) |  |
| Number of visits (mean (SD)) | 4.52 (2.49) | 5.25 (2.32) | < 0.001 |
| Participant characteristics for the Trails B times analysis |  |  |  |
| N (%) | 1339 (83.5) | 265 (16.5) |  |
| Trails B time (median [IQR]) | 60.00 [47.00, 80.00] | 68.00 [55.00, 90.00] | < 0.001 |
| Age (last visit) (mean (SD)) | 65.9 (9.12) | 69.84 (8.06) | < 0.001 |
| Sex (%) |  |  | .015 |
| F | 922 (68.9) | 203 (76.6) |  |
| M | 417 (31.1) | 62 (23.4) |  |
| ApoE4 status (%) |  |  | 0.001 |
| 0 ε4 alleles | 787 (58.8) | 184 (69.4) |  |
| 1 or 2 ε4 alleles | 552 (41.2) | 81 (30.6) |  |
| Education, years (mean (SD)) | 16.03 (2.79) | 16.22 (2.91) | 0.328 |
| Race (%) |  |  | 0.199 |
| Black | 125 (9.3) | 22 (8.3) |  |
| Non White/Non Black | 32 (2.4) | 2 (0.8) |  |
| White | 1182 (88.3) | 241 (90.9) |  |
| Number of tests (mean (SD)) | 4.57 (2.31) | 5.18 (2.09) | < 0.001 |

This difference was significant (N=1776, FET, p-value=0.03). We then performed a logistic regression to control for age, sex, race, ApoE4 status, education, and number of visits in the study, and found the significance of VBA status to degrade in its explanation of diagnosis (N=1776, GLMperm, p-value= 0.15).

Moreover, there was a significant dependency between VBA and Amyloid positivity (N=539, GLMperm, p-value=0.013, see Table 10), with the probability of VBA+ given Amyloid negativity (A-) higher than that in the Amyloid positive group (*P* [*V BA* + |*A*−] = 0.24 vs *P* [*V BA* + |*A*+] = 0.16). No such dependency was observed between VBA and tau positivity (N=509, FET, p-value=0.432). These facts suggest that the process by which a history of VBA leads to a positive diagnosis may be different from those that lead to an accumulation of amyloid and tau misfolded proteins in the brain. An additional fact of interest is that the VBA count is negatively correlated with ApoE4 positivity (N=1606, FET, p-value=0.005), with *P* [*V BA* +|*ApoE*4^+^] = 12.9% and *P* [*V BA* +|*ApoE*4*^−^*] = 18.9%. When correcting for age, sex and race, we found a negative coefficient for ApoE4 which was statistically significant (N=1606, GLMperm, p-value=0.002; see Table 19). ApoE4 positivity thus seems protective against VBA. Note that this effect was already observed on a restricted list of common infections [4], and is in line with the protective role described for ApoE4+ against viral and microbial infections (see Discussion).

We next considered the influence of VBAs on cognition. Both individual tests and composite tests were used. The time to complete the Trails B test was first used to assess cognition [29], as deemed more sensitive than, for example, the Trails A test. Significant differences were obtained for the time to complete the test across the different VBA levels, with a median of 60 seconds among VBA- participants and one of 68 seconds among VBA+ participants (see Figure 1, left). Controlling for covariates (age, sex, race, education, and number of tests taken) confirmed the positive significant effect of the VBA status on Trails B time (N=1604, FL, p-value=0.0042; see Suppl. Table 11). In addition to the Trails B test, the Stroop Test Color–Word and the Wechsler Adult Intelligence Scale–Revised Digit Symbol Total Score showed significant differences between VBA- and VBA+, with VBA+ participants doing worse (see Suppl. Table 7). The Preclinical Alzheimer’s Cognitive Composite (PACC): three-test version with Digit Symbol did not return significance, while the five-test version with CFL, Digit Symbol and Trails B tests did. The Executive Function composite score was also significantly lower among participants with VBA+ (N=1357, FL, p-value=1.97 × 10*^−^*^3^). Table 7 further indicates that the neuro-psychological scores associated with immediate learning and delayed recall did not show significant differences with VBA levels. Thus, VBA+ participants showed worse executive functions than VBA-, but not worse memory and learning scores. These results are qualitatively different from those obtained on the Amyloid positive participants in WRAP [18].

We next assessed whether the effect we observed on the Trails B time was specific to a subset of the VBA conditions used. A significant difference in the time to complete the Trails B test was observed for viral or dominantly viral infections (N = 1926, *N_V_* = 189 (9.81%), FL, p-value=0.001, see Table 12) and respiratory infections (*N_R_* = 157 (8.15%), FL, p-value=0.002, see Table 13), but not so for bacterial infections (*N_B_* = 119 (6.17%), WRST, p-value=0.47) or auto-immune conditions (*N_AI_* = 30 (1.56%), WRST, p-value=0.16).

### d.2 Protein Markers

Plasma proteins were next evaluated for their possible correlation to VBA level and cognitive status. A first insight into the concentrations of a small set of proteins was provided by the SIMOA assay (see Table 3 for details on each analyte). With respect to diagnosis, NFL, pTau-217, and GFAP all had significantly higher values among the C+ participants relative to the C- participants (Suppl. Table 8), as observed in other studies. Ab40 and Ab42 showed non-significant variations between the C- and C+ groups. The situation was different when we categorized participants by VBA. Ab42 showed significant variations across VBA groups, increasing from VBA- to VBA+ (N=1348, FL, p-value = 0.042). Ab40 also increased, but not significantly so. The observed increases in the remaining SIMOA markers were not significant between the VBA- and the VBA+ participants. This first series of comparisons thus revealed a discernible effect of VBA on plasma protein levels, which was different from that observed using cognitive diagnosis (C).

**Table 3:** Summary of plasma biomarkers for the SIMOA Analysis. Reported p values are computed using the Freedman-Lane permutation regression procedure [26] to control for covariates, after removal of outliers (*< Q*1 − 1.5 × *IQR* or *> Q*3 + 1.5 × *IQR*).

| Protein | VBA- |  | VBA+ |  | p-value |
| --- | --- | --- | --- | --- | --- |
|  | Median [IQR] | *N* | Median [IQR] | *N* |  |
| Ab40 | 96.3 [83.6, 113] | 1126 | 104 [91.1, 120] | 229 | 0.051 |
| Ab42 | 6.43 [5.37, 7.55] | 1121 | 6.92 [5.70, 7.92] | 227 | 0.041 |
| GFAP | 113 [76.9, 158] | 1103 | 122 [97.7, 168] | 222 | 0.904 |
| NFL | 15.5 [11.4, 21.6] | 1103 | 18.2 [13.6, 23.9] | 220 | 0.108 |
| pTau-217 | 0.309 [0.225, 0.455] | 1058 | 0.353 [0.26, 0.53] | 214 | 0.081 |

A larger set of proteins was next analyzed, which were quantified over a smaller set of participants (N=152). This analysis was performed in order to obtain more resolution on possible molecular mechanisms involved in the response to VBA in participants with evolving dementia. The analysis followed the NULISA immuno-based workflow [24]. The distributions and median values across VBA levels for the 28 markers we selected are shown in Table 4. As observed on the SIMOA dataset, neither NFL, nor pTau-217 or GFAP displayed significant variations across VBA levels under the NULISA protocol. Still, the difference in detected Ab42 levels remained significant between VBA- and VBA+ among the study participants. Among the markers specific to our NULISA set, a second one showed a significant variation between the VBA- and VBA+ groups, adhesion protein ICAM1 (see Table 4). While Ab42 levels were higher in VBA+ participants with respect to VBA- (increase of 19.6% of group median), ICAM1 amounts were lower among the VBA+ participants, with a median value down 12.9%. Both proteins were also controlled for covariates (see Table 16 and Table 17).

**Table 4:** Sample characteristics for the NULISA Analysis. The values for the selected proteins correspond to median and inter-quartile ranges for NPQs within a VBA level. p-values computed using WRST.

|  | VBA− | VBA+ | p |
| --- | --- | --- | --- |
| **Sample characteristics for the NULISA Analysis** |  |  |  |
| N (%) | 130 (85.5) | 22 (14.5) |  |
| Ab40 (median [IQR]) | 12.80 [12.47, 13.10] | 12.89 [12.65, 13.15] | 0.34 |
| Ab42 (median [IQR]) | 12.49 [12.20, 12.76] | 12.75 [12.50, 12.93] | **0.035** |
| ACHE (median [IQR]) | 13.15 [12.89, 13.39] | 13.21 [12.89, 13.32] | 0.88 |
| ANXA5 (median [IQR]) | 9.63 [8.43, 11.60] | 10.08 [8.87, 13.22] | 0.32 |
| APOE (median [IQR]) | 13.39 [13.13, 13.64] | 13.51 [13.32, 13.74] | 0.14 |
| BACE1 (median [IQR]) | 13.50 [13.36, 13.68] | 13.55 [13.41, 13.75] | 0.55 |
| CHI3L1 (median [IQR]) | 12.36 [11.77, 13.00] | 12.67 [12.01, 13.63] | 0.17 |
| CHIT1 (median [IQR]) | 13.69 [13.10, 14.24] | 13.82 [13.05, 14.42] | 0.44 |
| CNTN2 (median [IQR]) | 13.82 [13.26, 14.29] | 14.08 [13.48, 14.51] | 0.27 |
| FABP3 (median [IQR]) | 13.26 [13.01, 13.61] | 13.41 [13.16, 13.79] | 0.22 |
| GFAP (median [IQR]) | 15.03 [14.66, 15.46] | 15.22 [14.68, 15.37] | 0.38 |
| ICAM1 (median [IQR]) | 12.86 [12.56, 13.07] | 12.66 [12.29, 12.93] | **0.033** |
| IFNG (median [IQR]) | 10.93 [10.50, 11.51] | 11.07 [10.72, 11.57] | 0.31 |
| IL2 (median [IQR]) | 12.56 [12.29, 12.96] | 12.91 [12.41, 13.23] | 0.17 |
| IL7 (median [IQR]) | 13.24 [12.70, 13.72] | 13.16 [12.70, 13.52] | 0.42 |
| IL33 (median [IQR]) | 12.55 [12.26, 12.83] | 12.69 [12.34, 12.84] | 0.42 |
| NFL (median [IQR]) | 14.67 [14.27, 15.03] | 14.65 [14.48, 15.05] | 0.32 |
| NPTX2 (median [IQR]) | 13.46 [13.19, 13.70] | 13.62 [13.26, 13.70] | 0.43 |
| NRGN (median [IQR]) | 13.38 [11.74, 15.49] | 14.05 [12.59, 16.60] | 0.41 |
| pTau-181 (median [IQR]) | 13.28 [12.78, 13.72] | 13.37 [13.06, 13.69] | 0.79 |
| pTau-217 (median [IQR]) | 11.68 [10.89, 12.33] | 11.77 [10.88, 12.08] | 0.75 |
| pTau-231 (median [IQR]) | 13.63 [13.07, 14.21] | 13.78 [13.22, 14.23] | 0.69 |
| SNCA (median [IQR]) | 12.68 [12.14, 13.59] | 12.84 [12.22, 14.28] | 0.68 |
| TIMP3 (median [IQR]) | 15.40 [14.35, 16.62] | 15.37 [14.48, 16.17] | 0.63 |
| TNF (median [IQR]) | 13.18 [12.94, 13.44] | 13.29 [12.99, 13.66] | 0.23 |
| TREM2 (median [IQR]) | 12.35 [11.91, 12.76] | 12.23 [11.92, 12.66] | 0.57 |
| VCAM1 (median [IQR]) | 13.35 [13.16, 13.58] | 13.31 [13.12, 13.57] | 0.87 |
| VEGFD (median [IQR]) | 12.02 [11.73, 12.31] | 12.07 [11.81, 12.37] | 0.48 |
| Age (last visit) (mean (SD)) | 70.27 (7.15) | 72.37 (6.47) | 0.199 |
| Sex (%) |  |  | 0.309 |
| F | 89 (68.5) | 18 (81.8) |  |
| M | 41 (31.5) | 4 (18.2) |  |
| ApoE4 status (%) |  |  | 0.092 |
| 0 ε4 alleles | 59 (46.1) | 15 (68.2) | |
| 1 or 2 ε4 alleles | 69 (53.9) | 7 (31.8) | |
| Education, years (mean (SD)) | 16.75 (2.66) | 17.09 (3.57) | 0.604 |
| Race (%) |  |  | 0.305 |
| Black | 7 (5.4) | 3 (13.6) |  |
| Non White/Non Black | 2 (1.5) | 0 (0.0) |  |
| White | 121 (93.1) | 19 (86.4) |  |

Based on these results, we next assessed the relevance of combining these two markers to obtain a more robust VBA marker. The two markers were combined as per their direction of variation between the VBA- and VBA+ infection groups, using {*NPQ*(*Ab*42) −*NPQ*(*ICAM* 1)}. Since the NPQ values reported by NULISA are in log2 units, and 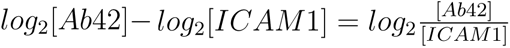, this corresponds mathematically to {Ab42/ICAM1}. As seen in Figure 3, combining these two proteins yielded a stronger infection marker (N=152, WRST, p-value=0.012). In a regression analysis with covariates, Ab42/ICAM1 was also significant (N=152, FL, p-value=0.003, see Table 18). The combined Ab42/ICAM1 marker proved altogether more robust than each protein considered individually (see values in Table 4). It should be noted that this combined marker also showed significantly different values across the diagnosis groups, with a decrease between C- and C+ (N=152, FL, p-value=0.02; see Discussion, Table 24).

### d.3 Correlations between protein markers

In order to detect further possible protein-protein relationships within the NULISA data, we next considered to what extent our protein markers were correlated to the Ab42/ICAM1 marker. The strongest correlations were observed for proteins Ab40, CHIT1, SOD1, CHI3L1, FABP3, pTau-217 and ApoE (N=152, SCT, p *<* 5×10*^−^*^2^). Among these markers, only Ab40 remained significant after multiple-testing correction (p *<* 5 × 10*^−^*^2^). When considering only VBA-particiants, proteins Ab40, FABP3, CHIT1 and IL33 were significantly correlated to Ab42/ICAM1, with again only Ab40 being still significant after multiple-testing correction (see Figure 6). Correlations among VBA+ participants returned significant p-values for proteins Ab40 (positive correlation) and ACHE (negative correlation, *i.e.* ICAM1-like behavior), with neither remaining significant after multiple-testing correction. This analysis, therefore, highlighted different protein-to-marker correlations across the VBA levels, with Ab40 consistently positively correlated to Ab42/ICAM1. Still, incorporating Ab40 to the combined marker did not improve the significance of the marker. This analysis nevertheless evidenced that protein ACHE could be an interesting second-degree marker among VBA+ participants.

## e Discussion

Based on our list of participant-reported infections and health conditions, we highlighted significant correlations between VBA and executive function. A correlation between VBA and a diagnosis of declining or impaired cognition was also observed, but lost significance after correcting for covariates (Table 9).

The relationship of VBA with the trails B test did not seem to be specific to some subset of our VBA list, since splitting our list into *e.g.* viral or respiratory conditions did maintain significant correlations, even after correcting for covariates (see Tables 12, 13). This suggests that it is not one specific VBA that affected cognition within our cohort, but that an overall immune impact is present. Regarding the cognitive tests which were most affected by a VBA+ status, tests related to executive functions were the most significantly affected by this status. In the literature regarding Alzheimer’s disease, executive function has been described as less affected than memory functions, while executive functions are affected in similar manners among subjects with cerebrovascular disease [30, 31].

By analyzing protein data among the study participants, we could highlight some correlations between VBAs and plasma protein amounts. Within the larger cohort used to analyze a restricted set of proteins (SIMOA data), we evidenced significant differences in protein amounts with VBA levels. While multiple markers (GFAP, NFL, pTau-217) showed significant variations with respect to cognitive diagnosis (Figure 2, left), only Ab42 had its distribution significantly affected by the VBA level (Figure 2, right). Ab42 also varied with the VBA level under the NULISA protocol, with significantly higher amounts in VBA+ participants (Table 4). Such an increase in Ab42 had already been described, both among healthy participants and among ADRD patients, using ELISA detection in serum samples [16].

**Figure 2:**
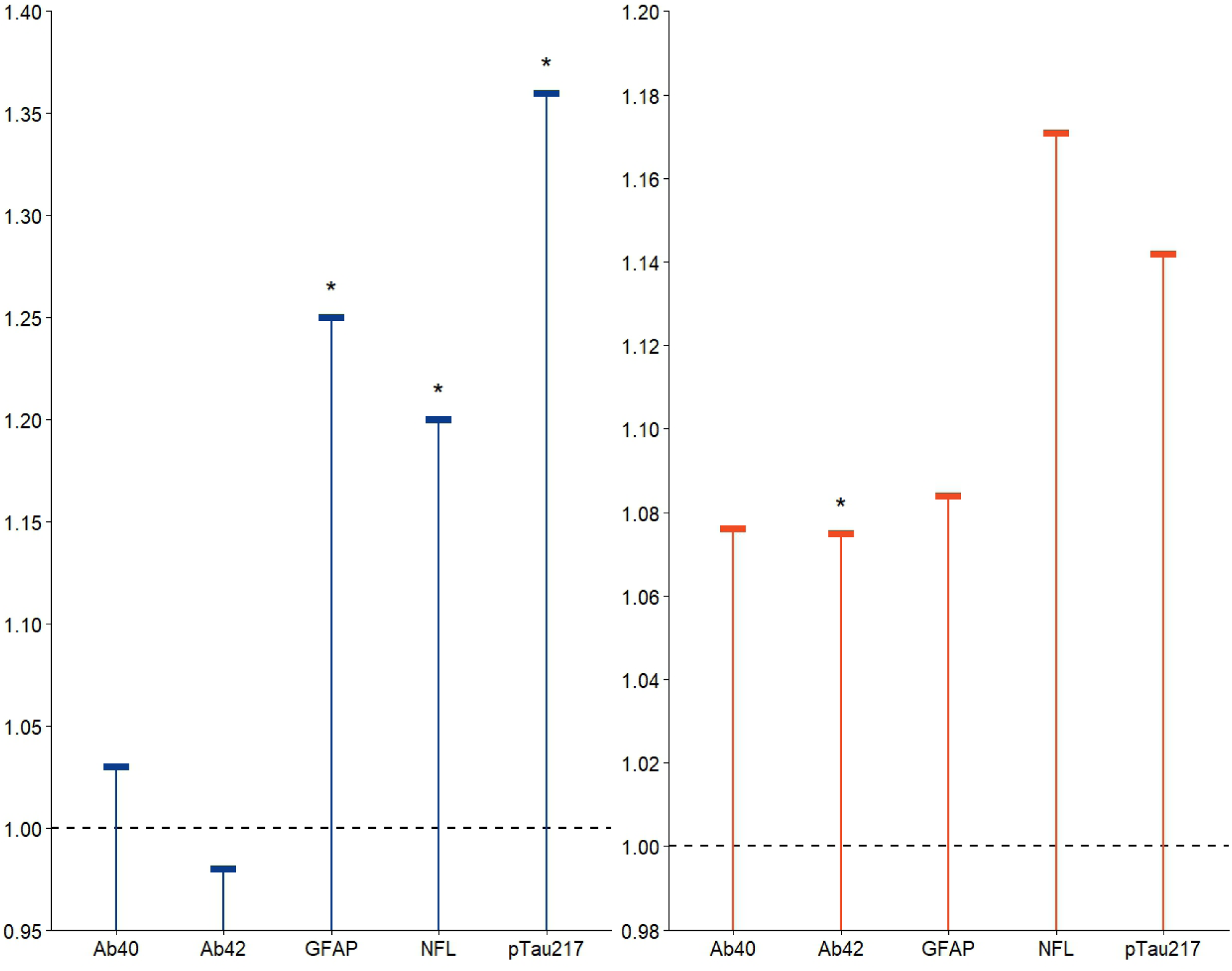
Ratios of protein amounts between the C+ and C-participants (left, blue) and the VBA+ and the VBA- participants (right, red) for the SIMOA markers. *: p *<* 5 × 10*^−^*^2^, p-values obtained using FL.

**Figure 3:**
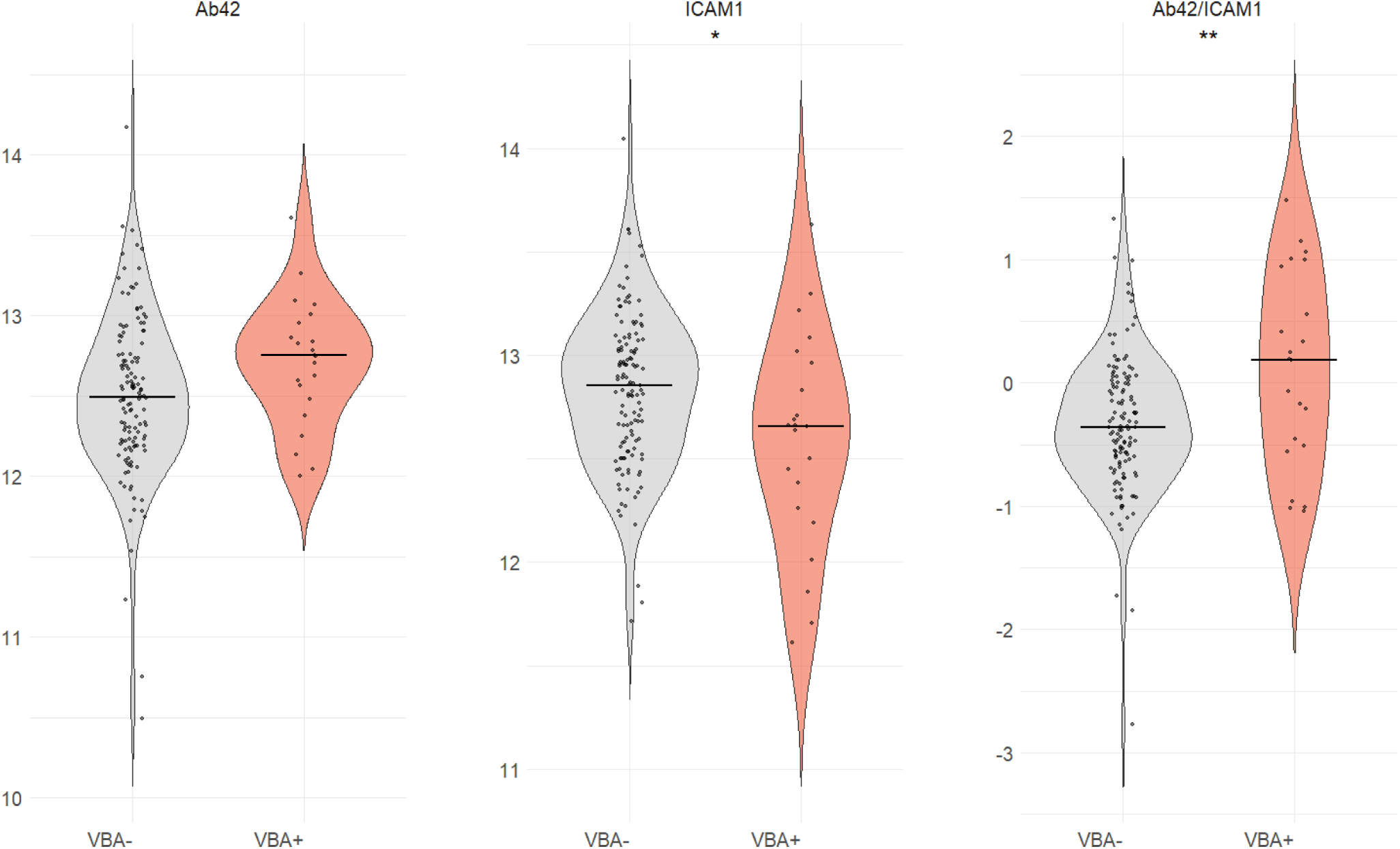
Distribution of NULISA markers per VBA level, grey: VBA-, light red: VBA+. Left, Ab42; middle, ICAM1; right, Ab42/ICAM1. ∗: *p <* 5×10*^−^*^2^, ∗∗: *p <* 5×10*^−^*^3^, p-values obtained using FL (see respective regression data in Supplementary material).

A novel plasma marker combining two single-protein markers was highlighted by our analysis: Ab42/ICAM1. It was significantly correlated both to VBA levels and to diagnosis. Previous studies found no significant variations of ICAM1 with dementia diagnosis (Chen and coworkers [32] evidenced a relation of adhesion proteins ALCAM and VCAM1, but not ICAM1, with cognitive impairment and age). Increased resolution in our analysis is possibly due to the quality of the detection under the NULISA approach. ICAM1 is a critical component at the contact site between the T-cell and the antigen-presenting cell [33]. BILF1 is a G-Protein Coupled Receptor expressed during the lytic replication cycle of EBV [34]. It has been shown to down-regulate the level of Class I Major Histocompatibility Complex at the surface of T-cells, thus enabling EBV to evade immune defenses, in a mechanism provoking an increase of NF-*κ*B and ICAM1 in Raji Cells [35]. ICAM1 was moreover shown to protect neurons from *Aβ* deposition and improve cognitive behavior in 5xFAD mice [36]. ICAM1 thus overall plays a protective role.

Ab42/ICAM1 seems to be an interesting early marker, as higher in {C-,VBA+} with respect to {C-,VBA-} participants (N=113, FL, p=0.015) and lower in C+ with respect to {C-,VBA+} participants (N=53, FL, p=0.066, see Table 25). In addition, the trails B analysis revealed significantly decreasing cognition following {C-,VBA-} →− {C-,VBA+} →− {C+} (see Figure 4; see WRST and FL p-values in Table 26). Hence, a decrease in Ab42/ICAM1 after its increase could possibly be related to a diagnosis evolving from C- to C+. This will be further explored using longitudinal NULISA data.

**Figure 4:**
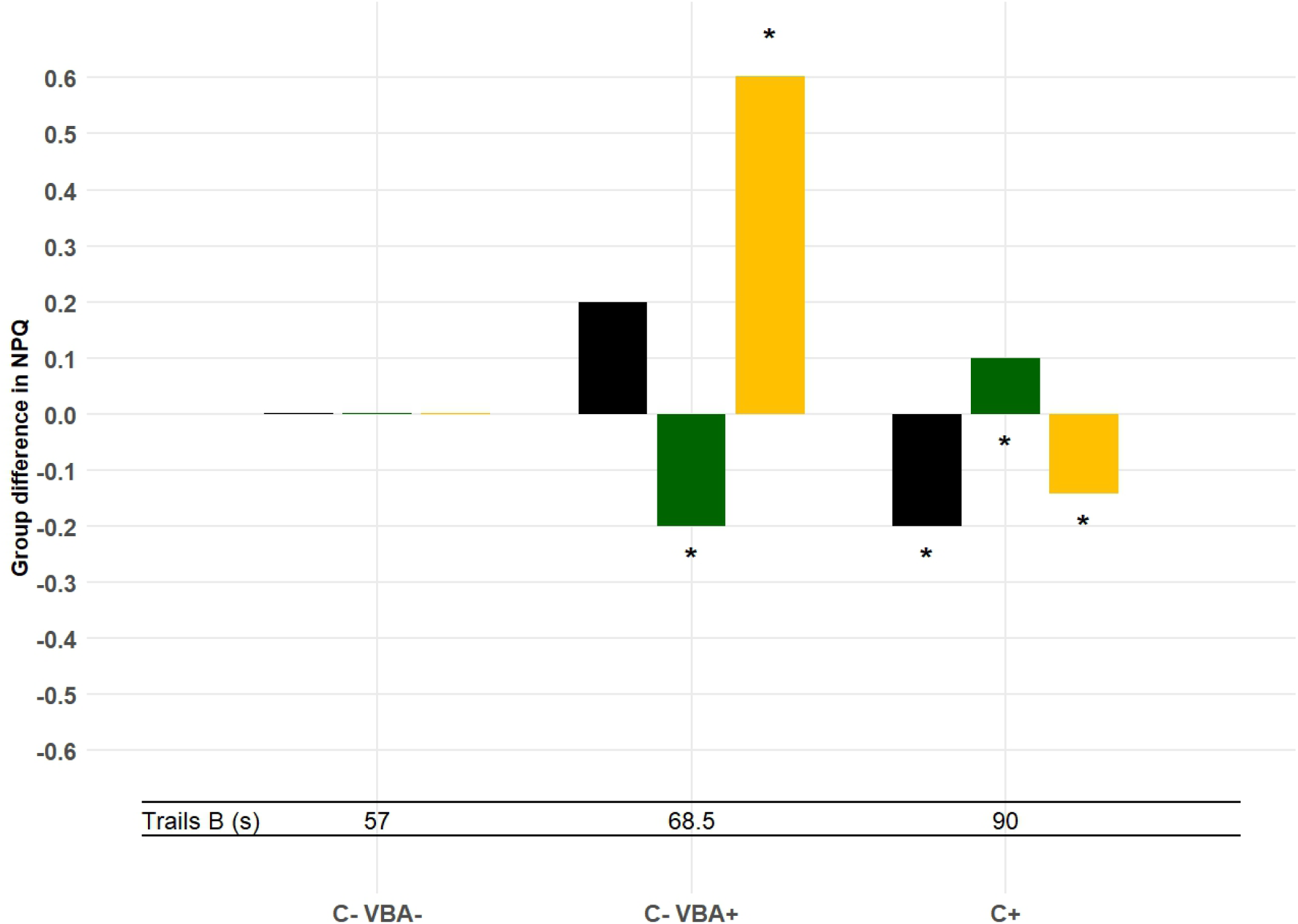
Differences in median values for Ab42 (black), ICAM1 (green) and Ab42/ICAM1 (yellow) as compared to the C- VBA- group (left, reference). *: p-values *<* 0.05 compared with the group immediately to the left. Regression analysis was used to adjust for covariates. Median Trails B times are also displayed.

We evidenced a smaller proportion of ApoE4 carriers among VBA+ participants than among VBA- (see Table 2). This result is consistent with the protective effect of ApoE4 already described on hepatitis C [37] and *in vitro* on *Plasmodium falciparum* [38]. Since ApoE4 is a known risk factor for AD [39], this result further supports the idea that the infection-related and A/T/N developments of ADRD [40] involve distinct physiological mechanisms.

Our list of VBAs included both infections and auto-immune conditions. Arguments in favor of an involvement of autoimmune conditions in dementia have been accumulating over the past decade, with an intensification over the past couple of years. Li and co-workers analyzed the records of hospitalized patients across Sweden over more than four decades [41]. Their analysis revealed a significantly higher standard incidence ratio (SIR) of dementia for patients diagnosed with some of the autoimmune conditions reported in WRAP (see Table 1). These included Grave’s disease (SIR=1.33), Hashimoto’s disease (SIR=1.44) and Sjögren’s syndrome (SIR=1.88). These authors could nevertheless not detect a significantly different incidence of dementia for patients diagnosed with Guillain-Barré’s syndrome (SIR=1.01) or lupus (SIR=1.01), both present in our list. This may be due to less representative samples, with only 18 and 17 patients respectively presenting these auto-immune conditions in their study.

Note that the infections and medical conditions that were used to characterize the WRAP participants in our study were based on a medical questionnaire. This presented limitations in the medical conditions which were accounted for in our analysis, with those presenting no or only mild symptoms most likely not accounted for. Discrepancy with population health data indeed exists for the following viral infections: HSV1 is known to have an overall prevalence of over 50% [42], while only 10 participants (less than 1 %) were documented as affected by this virus in our cohort. For CMV, reported prevalence levels range from around 35% in Canadians at age 40 [43], to 89% among an aged Black American cohort [44], while no participant reported an infection by CMV in our cohort (Table 1). This suggests that finer medical information regarding participants included in the cohort would enable one to detect more elaborate correlations between VBA on the one hand and their cognition and dementia status on the other hand.

In conclusion, after extracting medical information on infections and autoimmune diseases and correlating them with cognitive tests in the WRAP cohort, we found that participants with a history of at least one of these conditions (VBA+) had significantly lower executive function performance at their latest visit. Turning to plasma protein counts with the SIMOA protocol, we found higher levels of Ab42 in VBA+ compared to VBA-. Finally, using the NULISA protocol, we identified the ratio of Ab42 to ICAM1 to be significantly higher in VBA+ than in VBA- and a biomarker candidate for a history of VBA conditions.

## Data Availability

The data can be requested from the Wisconsin Registry for Alzheimer's Prevention at https;//wrap.wisc.edu/data-requests-2/

https://wrap.wisc.edu/data-requests-2/

## g. Acknowledgements/Conflicts/Funding Sources

This work was partly funded by the National Institute of Health RO1AG021155, R01EY032284, R01AG027161, and National Science Foundation #2136228. The authors thank Dr. Jacob Raber, Oregon Health and Science University, and Dr. Kirk Hogan, University of Wisconsin, for insightful discussions during this study, as well as Dr. Erin M. Jonaitis and Dr. Jessica Kirkland Caldwell for their comments.

---

**Table 5:**
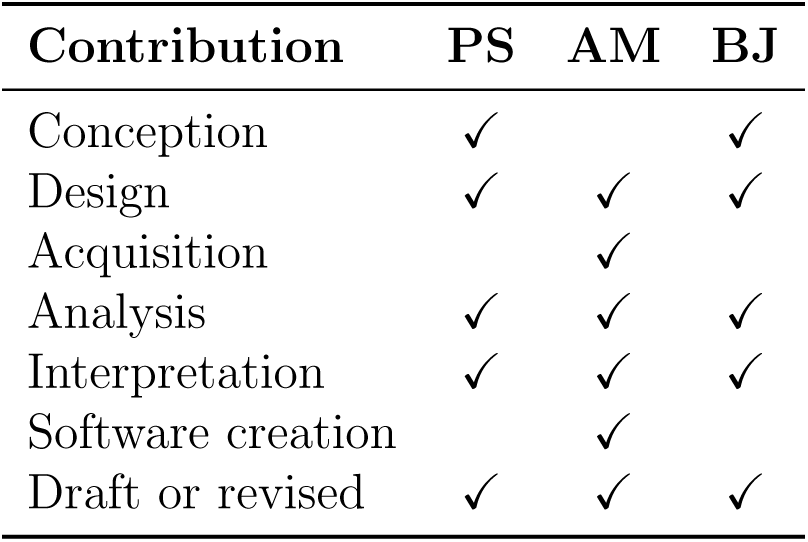
Contributions. PS: Patrick S. Slama, AM: Adam R. Macbale, BJ: Bruno M. Jedynak.

| Contribution | PS | AM | BJ |
| --- | --- | --- | --- |
| Conception | ✓ |  | ✓ |
| Design | ✓ | ✓ | ✓ |
| Acquisition |  | ✓ |  |
| Analysis | ✓ | ✓ | ✓ |
| Interpretation | ✓ | ✓ | ✓ |
| Software creation |  | ✓ |  |
| Draft or revised | ✓ | ✓ | ✓ |

## i. Supplementary material

**Figure 5:**
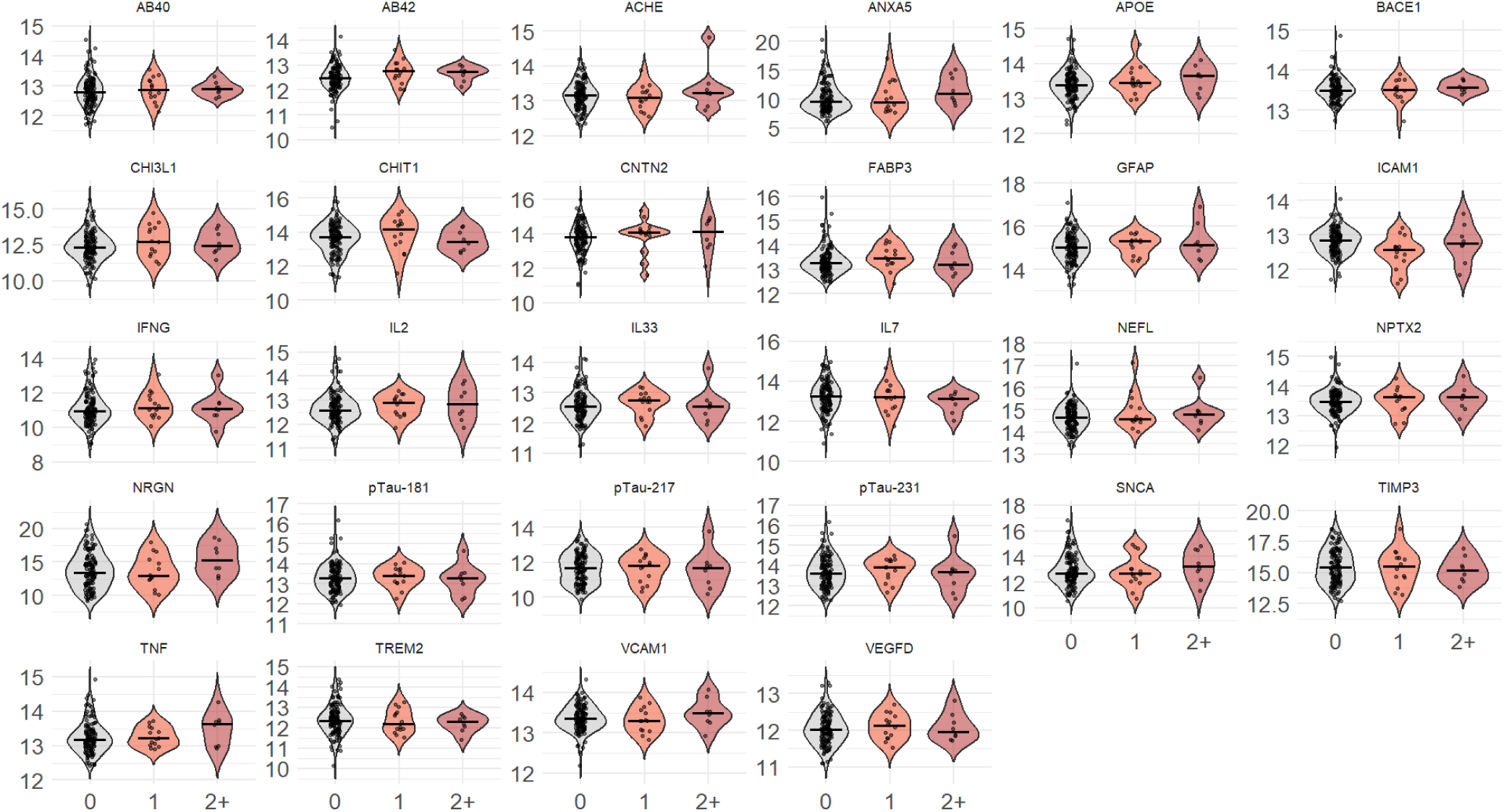
Distribution of the NPQ values for the selected 28 NULISA markers. Values are grouped per number of reported VBA.

**Figure 6:**
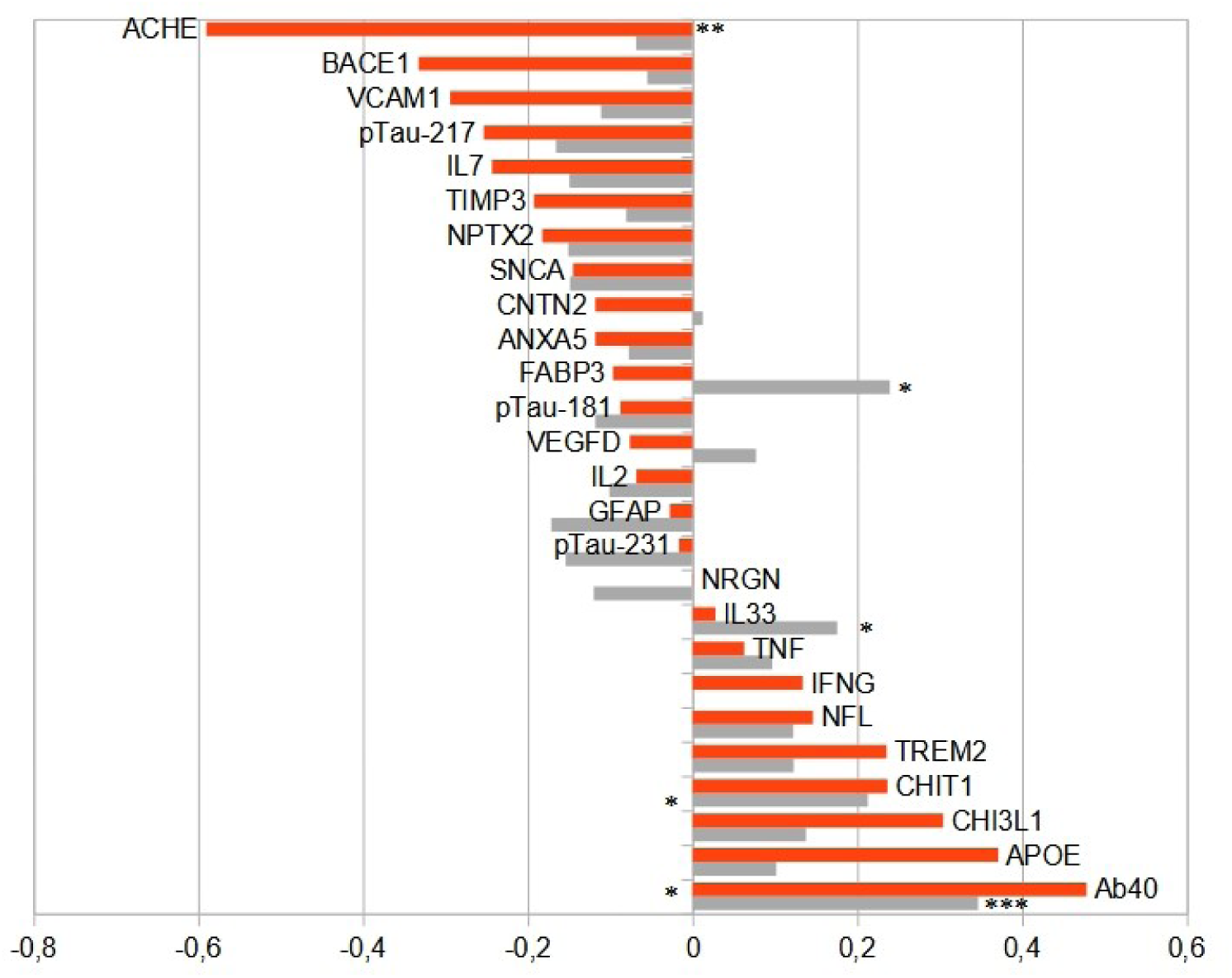
Correlations to marker Ab42/ICAM1 as a function of VBA level. Proteins are ordered by their *ρ*(*V BA*+) values. Grey bars: *ρ*(*V BA*−), red bars: *ρ*(*V BA*+). ∗: *p*(*ρ*) *<* 5 × 10*^−^*^2^, ∗∗: *p*(*ρ*) *<* 10*^−^*^3^, ∗ ∗ ∗: *p*(*ρ*) *<* 5 × 10*^−^*^2^ after multiple testing correction.

**Table 6:**
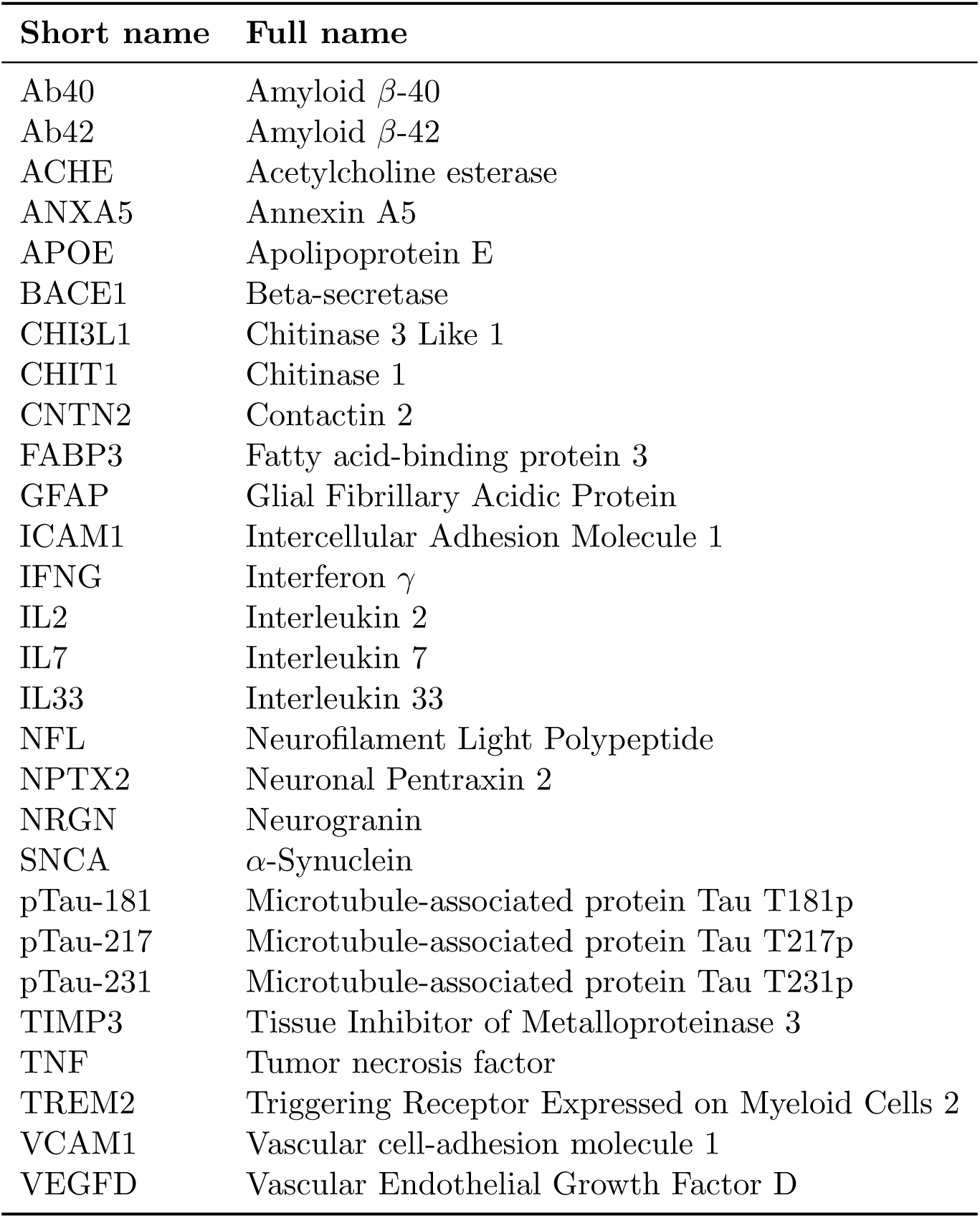
Abbreviated and full names of selected NULISA proteins included in our analysis.

| Short name | Full name |
| --- | --- |
| Ab40 | Amyloid β-40 |
| Ab42 | Amyloid β-42 |
| ACHE | Acetylcholine esterase |
| ANXA5 | Annexin A5 |
| APOE | Apolipoprotein E |
| BACE1 | Beta-secretase |
| CHI3L1 | Chitinase 3 Like 1 |
| CHIT1 | Chitinase 1 |
| CNTN2 | Contactin 2 |
| FABP3 | Fatty acid-binding protein 3 |
| GFAP | Glial Fibrillary Acidic Protein |
| ICAM1 | Intercellular Adhesion Molecule 1 |
| IFNG | Interferon $\gamma$ |
| IL2 | Interleukin 2 |
| IL7 | Interleukin 7 |
| IL33 | Interleukin 33 |
| NFL | Neurofilament Light Polypeptide |
| NPTX2 | Neuronal Pentraxin 2 |
| NRGN | Neurogranin |
| SNCA | α-Synuclein |
| pTau-181 | Microtubule-associated protein Tau T181p |
| pTau-217 | Microtubule-associated protein Tau T217p |
| pTau-231 | Microtubule-associated protein Tau T231p |
| TIMP3 | Tissue Inhibitor of Metalloproteinase 3 |
| TNF | Tumor necrosis factor |
| TREM2 | Triggering Receptor Expressed on Myeloid Cells 2 |
| VCAM1 | Vascular cell-adhesion molecule 1 |
| VEGFD | Vascular Endothelial Growth Factor D |

**Table 7:** Comparison of cognitive tests performances across VBA levels (p-values: WRST; adj. p-values obtained using regression analysis). **Lotraw:** Rey Auditory-Verbal Learning Test, Trials 1-5 Total Score **MmseTot:** Mini-Mental State Examination, Total Score **WMSrAr:** Wechsler Memory Scale-Revised Logical Memory I Total Score **BVMtTot:** Brief Visuospatial Memory Test-Revised Immediate Recall **Avltdr:** Rey Auditory-Verbal Learning Test Delayed Recall **WMSrTot2:** Wechsler Memory Scale-Revised Logical Memory II Total Score **BVMtDel:** Brief Visuospatial Memory Test-Revised Delayed Recall **StroopCW_Cor:** Stroop Test Color-Word (Corrected) **TrailsB:** Trail-Making Test Part B, time to complete. **WAISrTot:** Wechsler Adult Intelligence Scale-Revised Digit Symbol Total Score. **Exec_fn:** Executive function composite score (Trail-Making Test, Part B, Stroop Test, Color-Word (Corrected), Wechsler Adult Intelligence Scale-R, Digit Symbol Total Score) **PACC3_DS:** Preclinical Alzheimer’s Cognitive Composite (three-test version with Digit Symbol): Rey Auditory-Verbal Learning Test, Trials 1-5 Total Score, Wechsler Memory Scale-Revised, Logical Memory II Total Score, Wechsler Adult Intelligence Scale-R Digit Symbol Total Score **PACC5_CFL:** Preclinical Alzheimer’s Cognitive Composite (five-test version with CFL, Digit Symbol, and Trails B)

| Cognitive Test | VBA− |  | VBA+ |  | p-value | Adj. p-value |
| --- | --- | --- | --- | --- | --- | --- |
|  | Median | *N* | Median | *N* |  |  |
| Lotraw | 20 | 91 | 20 | 29 | 0.209 | 0.246 |
| MmseTot | 30 | 1268 | 29 | 265 | 0.555 | 0.936 |
| WMSrAr | 13 | 1313 | 14 | 268 | 0.487 | 0.505 |
| BVMtTot | 26 | 1243 | 26 | 263 | 0.273 | 0.0578 |
| Avltdr | 106 | 85 | 111 | 28 | 0.363 | 0.884 |
| WMSrTot2 | 23 | 1311 | 24 | 268 | 0.509 | 0.579 |
| BVMtDel | 10 | 1242 | 10 | 263 | 0.341 | 0.0940 |
| StroopCW_Cor | 104 | 1306 | 98 | 264 | $9.4 \times 10^{-5}$ | **0.0034** |
| TrailsB | 62 | 1447 | 69 | 278 | $9.8 \times 10^{-6}$ | **0.0042** |
| WAISrTot | 51 | 1261 | 48.5 | 264 | $1.4 \times 10^{-3}$ | **0.017** |
| Exec_fn | -0.121 | 1115 | -0.544 | 242 | $2.7 \times 10^{-5}$ | **0.0034** |
| PACC3_DS | -0.496 | 1260 | -0.614 | 263 | 0.165 | 0.183 |
| PACC5_CFL | -0.362 | 1251 | -0.547 | 260 | 0.0168 | **0.0386** |

**Table 8:** Summary of plasma biomarkers for the SIMOA Analysis as a function of cognitive status. Reported p values are computed using the Freedman-Lane permutation regression procedure to control for covariates, after removal of outliers (*< Q*1 − 1.5 × *IQR* or *> Q*3 + 1.5 × *IQR*).

| Protein | C- |  | C+ |  | p-value |
| --- | --- | --- | --- | --- | --- |
|  | Median [IQR] | *N* | Median [IQR] | *N* |  |
| Ab40 | 97.3 [84.4, 115] | 1033 | 100 [84.5, 118] | 347 | 0.38 |
| Ab42 | 6.53 [5.47, 7.78] | 1033 | 6.41 [5.25, 7.54] | 347 | 0.557 |
| GFAP | 113 [79.6, 159] | 1032 | 141 [93.9, 206] | 347 | $4 \times 10^{-4}$ |
| NFL | 15.5 [11.3, 22.0] | 1033 | 18.6 [14.3, 27.1] | 347 | $10^{-3}$ |
| pTau-217 | 0.311 [0.232, 0.476] | 1032 | 0.424 [0.270, 0.791] | 346 | $2 \times 10^{-4}$ |

**Table 9:** Logistic regression results predicting diagnosis. VBA status p-value computed by permutation of regression residuals.

| Variable | Estimate | Std. Error | z value | Pr(> \|z\|) |
| --- | --- | --- | --- | --- |
| (Intercept) | -5.600 | 0.671 | -8.35 | $< 2 \times 10^{-16}$ |
| VBA status | 0.243 | 0.160 | 1.52 | 0.15 |
| Age | 0.097 | 0.010 | 9.95 | $< 2 \times 10^{-16}$ |
| Sex = Male | 0.631 | 0.131 | 4.81 | $1.5 \times 10^{-6}$ |
| ApoE4 status | 0.567 | 0.127 | 4.46 | $8.1 \times 10^{-6}$ |
| Education (years) | -0.080 | 0.023 | -3.45 | $5.6 \times 10^{-4}$ |
| Race = Non White/Non Black | 0.066 | 0.479 | 0.14 | 0.891 |
| Race = White | -0.673 | 0.212 | -3.17 | $1.5 \times 10^{-3}$ |
| Number of visits | -0.142 | 0.036 | -3.93 | $8.6 \times 10^{-5}$ |

**Table 10:** Logistic regression results predicting VBA status as a function of amyloid positivity and covariates. Amyloid positivity p-value computed by permutation of regression residuals.

| Variable | Estimate | Std. Error | z value | Pr(> \|z\|) |
| --- | --- | --- | --- | --- |
| (Intercept) | -5.011 | 1.518 | -3.30 | 0.00096 |
| Amyloid positivity status | -0.692 | 0.286 | -2.41 | 0.01290 |
| Age | 0.051 | 0.019 | 2.71 | 0.00676 |
| Sex = Male | -0.384 | 0.267 | -1.44 | 0.15023 |
| ApoE4 status | -0.033 | 0.259 | -0.13 | 0.90002 |
| Education (years) | 0.046 | 0.041 | 1.12 | 0.26225 |
| Race = Non White/Non Black | -0.078 | 1.290 | -0.06 | 0.95183 |
| Race = White | -0.492 | 0.529 | -0.93 | 0.35255 |
| Number of visits | -0.002 | 0.081 | -0.03 | 0.97704 |

**Table 11:** Linear regression results predicting log(Trails B time). P-values computed by the Freedman-Lane permutation regression procedure.

| Variable | Estimate | Std. Error | t value | Pr(> \|t\|) |
| --- | --- | --- | --- | --- |
| (Intercept) | 3.568 | 0.091 | 39.08 | $< 2 \times 10^{-16}$ |
| VBA status | 0.074 | 0.025 | 2.93 | $4.2 \times 10^{-3}$ |
| Age | 0.020 | 0.001 | 16.20 | $2 \times 10^{-4}$ |
| Sex = Male | 0.098 | 0.020 | 4.83 | $2 \times 10^{-4}$ |
| ApoE4 status | 0.064 | 0.019 | 3.36 | $4 \times 10^{-4}$ |
| Education (years) | -0.018 | 0.003 | -5.52 | $2 \times 10^{-4}$ |
| Race = Non White/Non Black | -0.148 | 0.071 | -2.08 | 0.039 |
| Race = White | -0.367 | 0.035 | -10.58 | $2 \times 10^{-4}$ |
| Number of tests | -0.040 | 0.005 | -7.79 | $2 \times 10^{-4}$ |

**Table 12:** Linear regression results predicting log(Trails B time) as a function of viral (V) condition status and covariates. P-values computed by the Freedman-Lane permutation regression procedure.

| Variable | Estimate | Std. Error | t value | Pr(> \|t\|) |
| --- | --- | --- | --- | --- |
| (Intercept) | 3.513 | 0.074 | 47.34 | $< 2 \times 10^{-16}$ |
| V status | 0.108 | 0.031 | 3.52 | 0.0014 |
| Race = Non White/Non Black | -0.172 | 0.073 | -2.37 | 0.0190 |
| Race = White | -0.443 | 0.033 | -13.28 | 0.0002 |
| Age | 0.015 | 0.001 | 14.01 | 0.0002 |
| Sex = Male | 0.084 | 0.021 | 4.08 | 0.0002 |
| ApoE4 status | 0.063 | 0.019 | 3.25 | 0.0020 |

**Table 13:** Linear regression results predicting log(Trails B time) as a function of respiratory (R) condition status and covariates. P-values computed by the Freedman-Lane permutation regression procedure.

| Variable | Estimate | Std. Error | t value | Pr(> \|t\|) |
| --- | --- | --- | --- | --- |
| (Intercept) | 3.511 | 0.074 | 47.28 | $< 2 \times 10^{-16}$ |
| R status | 0.104 | 0.033 | 3.11 | 0.0020 |
| Race = Non White/Non Black | -0.168 | 0.073 | -2.31 | 0.0222 |
| Race = White | -0.444 | 0.033 | -13.27 | 0.0002 |
| Age | 0.015 | 0.001 | 14.10 | 0.0002 |
| Sex = Male | 0.081 | 0.021 | 3.94 | 0.0002 |
| ApoE4 status | 0.061 | 0.019 | 3.18 | 0.0020 |

**Table 14:** Linear regression results predicting SIMOA Ab42 levels. Reported p values are computed using the Freedman-Lane permutation regression procedure to control for covariates, after removal of outliers (*< Q*1 − 1.5 × *IQR* or *> Q*3 + 1.5 × *IQR*).

| Variable | Estimate | Std. Error | t value | Pr(> \|t\|) |
| --- | --- | --- | --- | --- |
| (Intercept) | 5.440 | 0.492 | 11.05 | $< 2 \times 10^{-16}$ |
| VBA status | 0.255 | 0.125 | 2.04 | 0.041 |
| Age | 0.016 | 0.006 | 2.57 | 0.009 |
| Sex = Male | -0.242 | 0.101 | -2.39 | 0.017 |
| ApoE4 status | -0.650 | 0.096 | -6.78 | 0.0002 |
| Education (years) | 0.003 | 0.017 | 0.17 | 0.861 |
| Race = Non White/Non Black | -0.510 | 0.497 | -1.03 | 0.313 |
| Race = White | 0.316 | 0.181 | 1.75 | 0.079 |

**Table 15:** Linear regression results predicting SIMOA Ab40 levels. Reported p values are computed using the Freedman-Lane permutation regression procedure to control for covariates, after removal of outliers (*< Q*1 − 1.5 × *IQR* or *> Q*3 + 1.5 × *IQR*).

| Variable | Estimate | Std. Error | t value | Pr(> \|t\|) |
| --- | --- | --- | --- | --- |
| (Intercept) | 57.493 | 6.324 | 9.09 | $< 2 \times 10^{-16}$ |
| VBA status | 3.098 | 1.600 | 1.94 | 0.051 |
| Age | 0.565 | 0.080 | 7.05 | 0.0002 |
| Sex = Male | -2.365 | 1.296 | -1.83 | 0.066 |
| ApoE4 status | -2.948 | 1.231 | -2.39 | 0.017 |
| Education (years) | 0.123 | 0.217 | 0.57 | 0.575 |
| Race = Non White/Non Black | -6.783 | 6.387 | -1.06 | 0.279 |
| Race = White | 4.664 | 2.310 | 2.02 | 0.045 |

**Table 16:** Linear regression results predicting NULISA Ab42 levels. Reported p values are computed using the Freedman–Lane permutation regression procedure to control for covariates.

| Variable | Estimate | Std. Error | t value | Pr(> \|t\|) |
| --- | --- | --- | --- | --- |
| (Intercept) | 13.072 | 0.529 | 24.72 | $< 2 \times 10^{-16}$ |
| VBA status | 0.213 | 0.119 | 1.79 | 0.069 |
| Age | -0.008 | 0.006 | -1.29 | 0.195 |
| Sex = Male | -0.061 | 0.092 | -0.67 | 0.504 |
| ApoE4 status | -0.123 | 0.085 | -1.45 | 0.154 |
| Education (years) | -0.010 | 0.015 | -0.65 | 0.510 |
| Race = Non White/Non Black | 0.045 | 0.533 | 0.08 | 0.915 |
| Race = White | 0.242 | 0.177 | 1.37 | 0.180 |

**Table 17:** Linear regression results predicting NULISA ICAM1 levels. Reported p values are computed using the Freedman–Lane permutation regression procedure to control for covariates.

| Variable | Estimate | Std. Error | t value | Pr(> \|t\|) |
| --- | --- | --- | --- | --- |
| (Intercept) | 12.903 | 0.420 | 30.74 | $< 2 \times 10^{-16}$ |
| VBA status | -0.218 | 0.095 | -2.30 | 0.025 |
| Age | 0.003 | 0.005 | 0.69 | 0.493 |
| Sex = Male | 0.094 | 0.073 | 1.28 | 0.203 |
| ApoE4 status | 0.040 | 0.067 | 0.59 | 0.561 |
| Education (years) | -0.019 | 0.012 | -1.60 | 0.109 |
| Race = Non White/Non Black | 0.405 | 0.423 | 0.96 | 0.308 |
| Race = White | -0.046 | 0.141 | -0.32 | 0.745 |

**Table 18:** Linear regression results predicting NULISA Ab42/ICAM1 levels. Reported p values are computed using the Freedman-Lane permutation regression procedure to control for covariates.

| Variable | Estimate | Std. Error | t value | Pr(> \|t\|) |
| --- | --- | --- | --- | --- |
| (Intercept) | 0.170 | 0.626 | 0.27 | 0.787 |
| VBA status | 0.431 | 0.141 | 3.05 | 0.003 |
| Age | -0.011 | 0.007 | -1.55 | 0.124 |
| Sex = Male | -0.155 | 0.109 | -1.43 | 0.158 |
| ApoE4 status | -0.163 | 0.100 | -1.62 | 0.103 |
| Education (years) | 0.009 | 0.018 | 0.52 | 0.593 |
| Race = Non White/Non Black | -0.360 | 0.631 | -0.57 | 0.52 |
| Race = White | 0.288 | 0.210 | 1.37 | 0.156 |

**Table 19:** Logistic regression results predicting VBA status as a function of ApoE4. ApoE4 status p-value computed by permutation of regression residuals.

| Variable | Estimate | Std. Error | z value | Pr(> \|z\|) |
| --- | --- | --- | --- | --- |
| (Intercept) | -4.875 | 0.622 | -7.83 | $4.81 \times 10^{-15}$ |
| ApoE4 status | -0.418 | 0.147 | -2.84 | 0.002 |
| Age | 0.053 | 0.009 | 6.11 | $9.71 \times 10^{-10}$ |
| Sex = Male | -0.465 | 0.159 | -2.92 | 0.00355 |
| Race = Non White/Non Black | -0.922 | 0.773 | -1.19 | 0.233 |
| Race = White | -0.094 | 0.245 | -0.38 | 0.702 |

**Table 20:** Linear regression results for WAIS–revised Digit Symbol Total Score. P-values computed using the Freedman–Lane permutation regression procedure.

| Variable | Estimate | Std. Error | t value | Pr(> \|t\|) |
| --- | --- | --- | --- | --- |
| (Intercept) | 78.639 | 3.043 | 25.85 | $< 2 \times 10^{-16}$ |
| VBA status | -1.709 | 0.731 | -2.34 | 0.0170 |
| Sex = Male | -5.904 | 0.607 | -9.73 | 0.0002 |
| Race = Non White/Non Black | 7.468 | 2.813 | 2.65 | 0.0106 |
| Race = White | 9.052 | 1.168 | 7.75 | 0.0002 |
| Education (years) | 0.518 | 0.100 | 5.17 | 0.0002 |
| Age | -0.641 | 0.041 | -15.77 | 0.0002 |
| ApoE4 status | -1.709 | 0.569 | -3.00 | 0.0034 |
| Number of tests | 0.420 | 0.163 | 2.58 | 0.0116 |

**Table 21:**
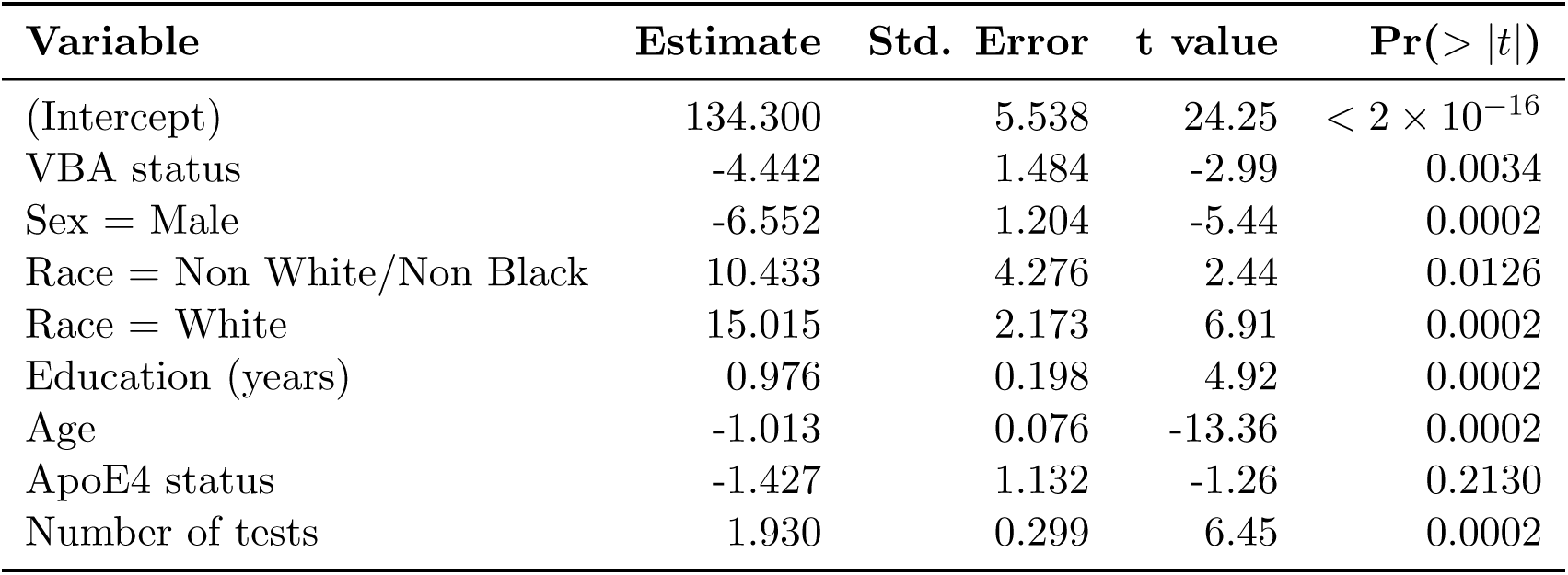
Linear regression results for Stroop Test Color–Word. P-values computed using the Freedman–Lane permutation regression procedure.

| Variable | Estimate | Std. Error | t value | Pr(> \|t\|) |
| --- | --- | --- | --- | --- |
| (Intercept) | 134.300 | 5.538 | 24.25 | $< 2 \times 10^{-16}$ |
| VBA status | -4.442 | 1.484 | -2.99 | 0.0034 |
| Sex = Male | -6.552 | 1.204 | -5.44 | 0.0002 |
| Race = Non White/Non Black | 10.433 | 4.276 | 2.44 | 0.0126 |
| Race = White | 15.015 | 2.173 | 6.91 | 0.0002 |
| Education (years) | 0.976 | 0.198 | 4.92 | 0.0002 |
| Age | -1.013 | 0.076 | -13.36 | 0.0002 |
| ApoE4 status | -1.427 | 1.132 | -1.26 | 0.2130 |
| Number of tests | 1.930 | 0.299 | 6.45 | 0.0002 |

**Table 22:** Linear regression results for PACC5 CFL. P-values computed using the Freedman– Lane permutation regression procedure.

| Variable | Estimate | Std. Error | t value | Pr(> \|t\|) |
| --- | --- | --- | --- | --- |
| (Intercept) | 1.283 | 0.326 | 3.94 | 0.0119 |
| VBA status | -0.158 | 0.079 | -2.01 | 0.0386 |
| Sex = Male | -0.666 | 0.065 | -10.23 | 0.0002 |
| Race = Non White/Non Black | 0.704 | 0.302 | 2.33 | 0.0246 |
| Race = White | 1.183 | 0.127 | 9.33 | 0.0002 |
| Education (years) | 0.098 | 0.011 | 9.12 | 0.0002 |
| Age | -0.067 | 0.004 | -15.65 | 0.0002 |
| ApoE4 status | -0.267 | 0.061 | -4.37 | 0.0002 |
| Number of tests | 0.120 | 0.019 | 6.36 | 0.0002 |

**Table 23:**
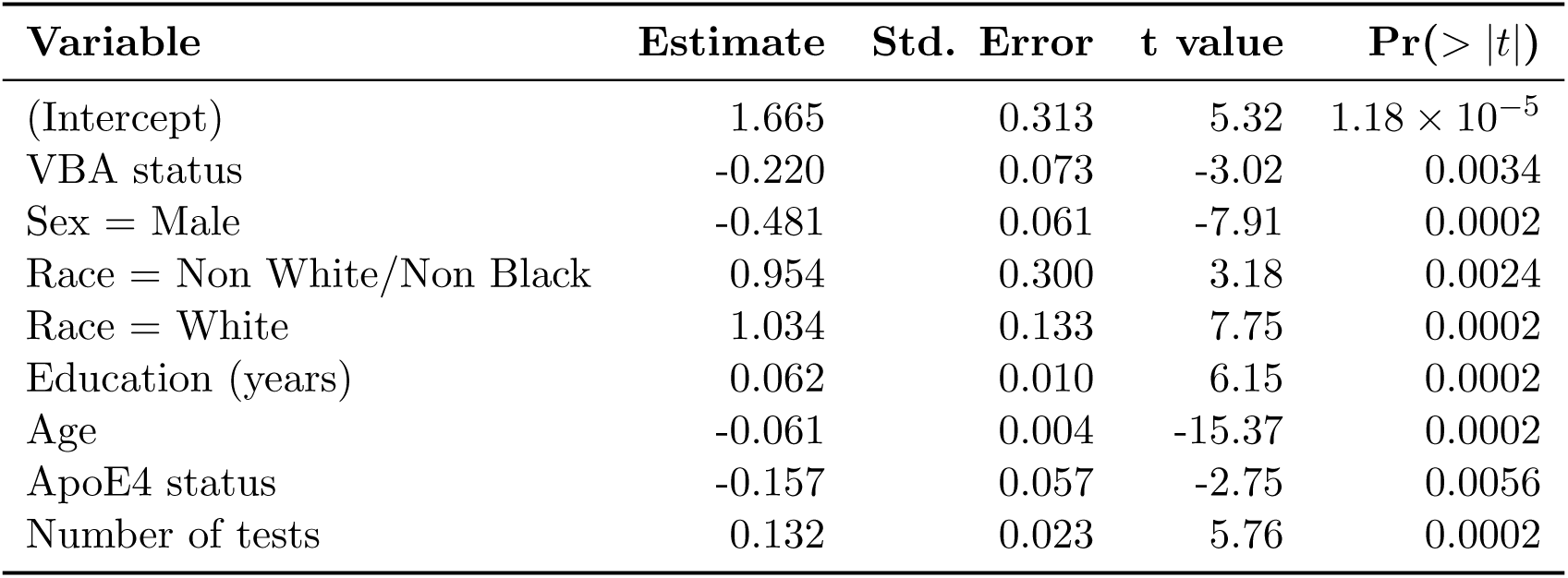
Linear regression results for Executive Function Composite. P-values computed using the Freedman–Lane permutation regression procedure.

**Table 24:**
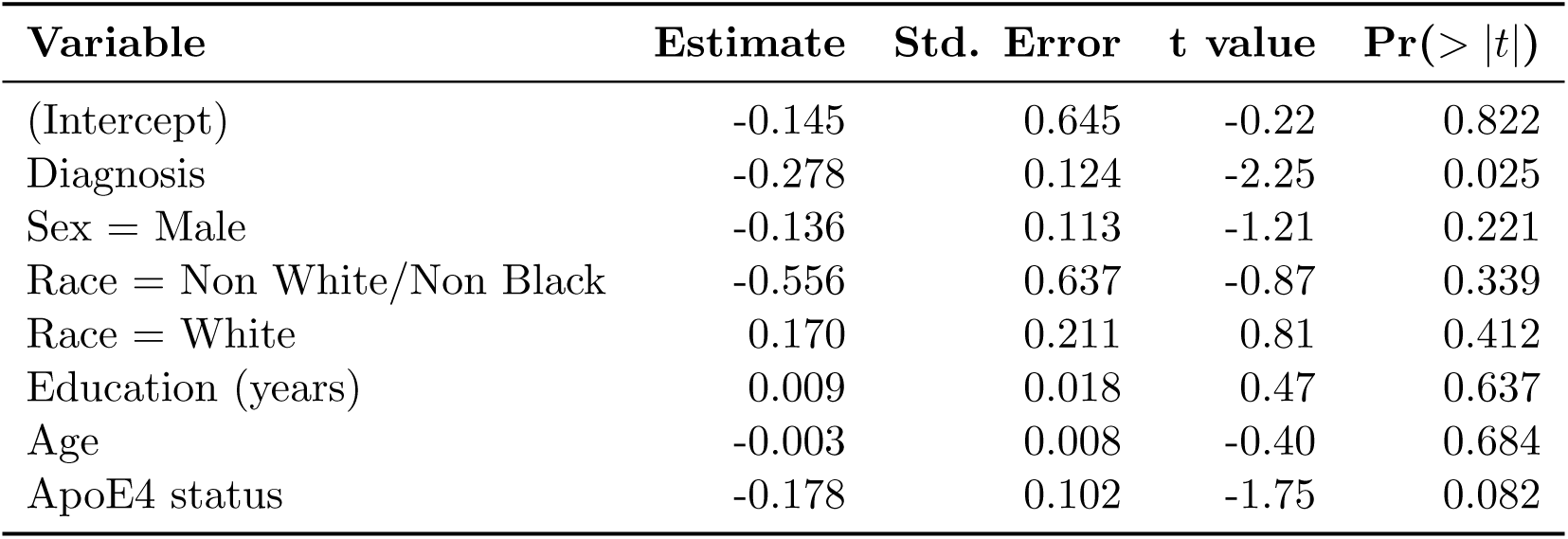
Linear regression results predicting Ab42/ICAM1 differences using diagnosis and covariates. Reported p values are computed using the Freedman–Lane permutation regression procedure.

| Variable | Estimate | Std. Error | t value | Pr(> \|t\|) |
| --- | --- | --- | --- | --- |
| (Intercept) | -0.145 | 0.645 | -0.22 | 0.822 |
| Diagnosis | -0.278 | 0.124 | -2.25 | 0.025 |
| Sex = Male | -0.136 | 0.113 | -1.21 | 0.221 |
| Race = Non White/Non Black | -0.556 | 0.637 | -0.87 | 0.339 |
| Race = White | 0.170 | 0.211 | 0.81 | 0.412 |
| Education (years) | 0.009 | 0.018 | 0.47 | 0.637 |
| Age | -0.003 | 0.008 | -0.40 | 0.684 |
| ApoE4 status | -0.178 | 0.102 | -1.75 | 0.082 |

**Table 25:**
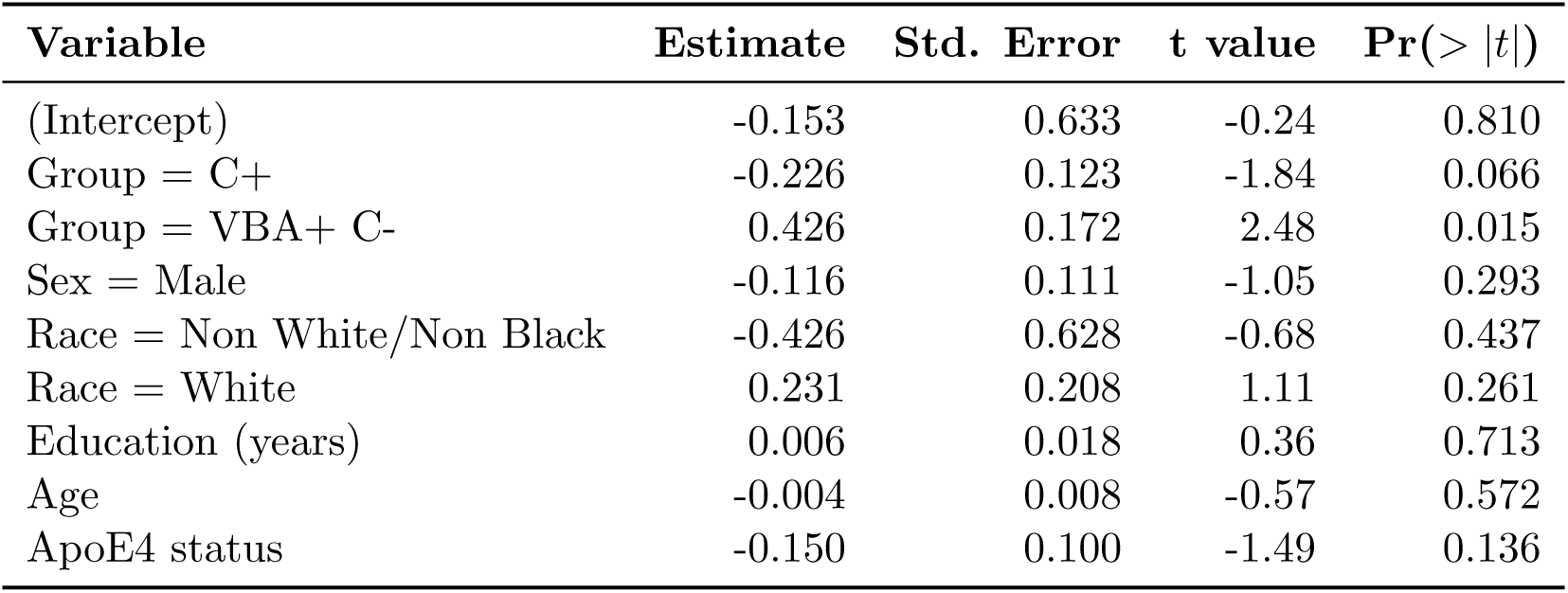
Linear regression results predicting Ab42/ICAM1 with VBA- C- as the reference group. Reported p values are computed using the Freedman–Lane permutation regression procedure.

| Variable | Estimate | Std. Error | t value | Pr(> \|t\|) |
| --- | --- | --- | --- | --- |
| (Intercept) | -0.153 | 0.633 | -0.24 | 0.810 |
| Group = C+ | -0.226 | 0.123 | -1.84 | 0.066 |
| Group = VBA+ C- | 0.426 | 0.172 | 2.48 | 0.015 |
| Sex = Male | -0.116 | 0.111 | -1.05 | 0.293 |
| Race = Non White/Non Black | -0.426 | 0.628 | -0.68 | 0.437 |
| Race = White | 0.231 | 0.208 | 1.11 | 0.261 |
| Education (years) | 0.006 | 0.018 | 0.36 | 0.713 |
| Age | -0.004 | 0.008 | -0.57 | 0.572 |
| ApoE4 status | -0.150 | 0.100 | -1.49 | 0.136 |

**Table 26:** Pairwise comparisons of Trails B completion time across C and VBA groups. Reported Trails B values are group medians. Wilcoxon rank-sum tests are shown alongside Freedman–Lane (FL) permutation regression *p*-values adjusting for covariates.

| **Comparison** | **Trails B (s)** | **Wilcoxon $p$** | **FL permutation $p$** |
| --- | --- | --- | --- |
| C- VBA- vs C- VBA+ | 57 vs 68.5 | $3.1 \times 10^{-9}$ | 0.0004 |
| C- VBA- vs C+ | 57 vs 90 | $< 2 \times 10^{-16}$ | 0.0002 |
| C- VBA+ vs C+ | 68.5 vs 90 | $4.3 \times 10^{-6}$ | 0.0002 |

